# COVID-19: Saving lives and livelihoods using population density driven testing

**DOI:** 10.1101/2020.06.28.20142174

**Authors:** Karim I. Budhwani, Henna Budhwani, Ben Podbielski

**Affiliations:** CerFlux, Inc., Birmingham, AL 35205; University of Alabama at Birmingham (UAB), Birmingham, AL 35294; Protective Life Corp., Birmingham, AL 35223

## Abstract

SARS-CoV-2 transmission risk generally increases with proximity of those shedding the virus to those susceptible to infection. Thus, this risk is a function of both number of people and the area which they occupy. However, the latter continues to evade COVID-19 testing policy. Increased testing in areas with lower population density, has the potential to induce a false sense of security even as cases continue to rise sharply overall.

## Policy Proposal

COVID-19 testing is typically measured per capita; globally tests and cases are reported per million while local authorities report these per 100,000 people.^1–3^ This approach is simple and generally well-accepted both in economic spheres and in healthcare research. However, this simplicity belies the underlying fallacy in applying per capita models to test transmission characteristics of SARS-CoV-2. The transmission risk profile for 20 people in an elevator is substantially different from that for 20 people spread across a football field; this was the fundamental premise for social-distancing and lockdowns to “flatten the curve.” In this two-part study, we analyze per capita COVID-19 testing data reported for Alabama to evaluate whether testing realignment along population density, rather than density agnostic per capita, would be more effective. Alabama is one of several states currently experiencing notable increases in new cases.

## Methods

Population characteristics and population density for all 67 Alabama counties were obtained from the 2018 American Community Survey (US Census Bureau). Numbers of tests administered and positive cases of COVID-19 are updated daily by the Alabama Department of Public Health. This data was obtained on May 18^th^ for initial assessment (Figure 1) and again on June 15^th^ for prospective analysis (Figure 2). Descriptive statistical analyses were performed to calculate the total number of tests per 100,000 people using county population as denominator, and subsequently dividing this by county population density, density squared, and square root of density as illustrative proxies^4,5^ of more complex population density test rate models. All study data were publicly available thereby obviating institutional review board approval.

## Results

Although we use Alabama for illustration, most states report statistics in this manner making our processes replicable in other states. The heatmap on the left appears to indicate widespread testing per 100,000 people^6^ by county. However, this does not reflect which areas are more like 20 people in an elevator vs. spread across a football field^7^ (Figure 1B). Overlaying the two (Figure 1C), provides a sense of magnitude by which we may be over-testing in areas with a natural spatial defense against transmission while severely under-testing in areas with elevated risk of transmission.

**Figure 1.**
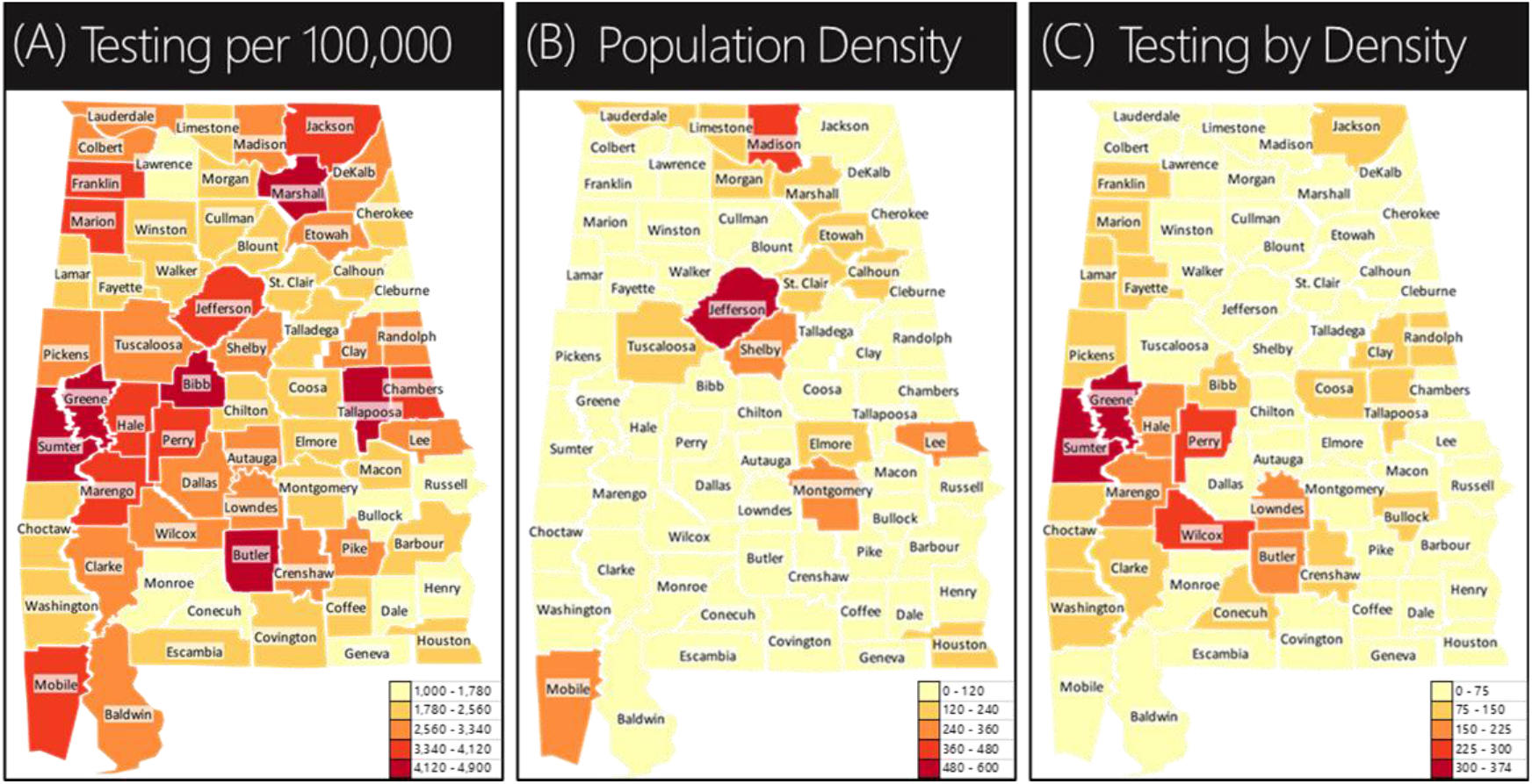
Per capita and population density heatmaps for COVID-19 tests between April 1 and May 18. (A) Heatmap of tests per 100,000 appear to indicate widespread testing across the state. (B) However, this fails to account for sparsely populated areas that may inherently provide spatial distancing. (C) Overlaying the two shows current testing by population density. Absent a population density driven testing approach, the risk of deriving a false sense of security is greater.

In the second part of the study, conducted during phased economic reengagement, data was collected to prospectively analyze distribution of tests and cases vis-à-vis population density. As anticipated^8^, new cases were disproportionately more prevalent in densely populated areas (Figure 2), despite relatively fewer tests per population density, suggesting that cases in these areas may be understated.

**Figure 2.**
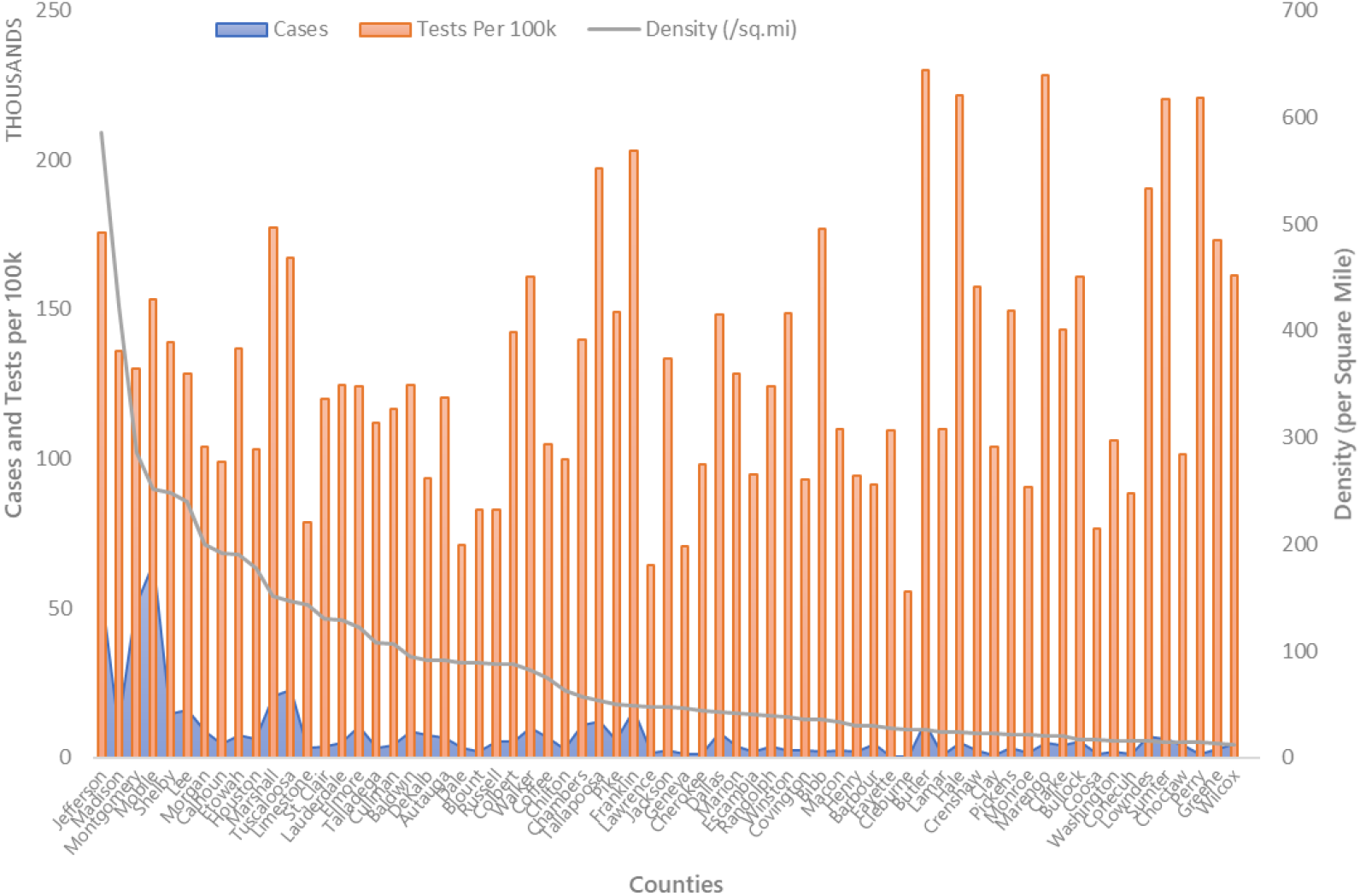
COVID-19 testing during phased reopening of the Alabama economy from May 18 to June 15. Tests reported per 100,000 during this period once again appeared to present an impression of widespread statewide testing. However, there was little correlation (r=0.28, p=0.02) between tests per capita and number of cases. In terms of population density, new cases were higher in areas with higher population density, despite relatively lower test rates as a function of density. This suggests that a population density driven testing strategy would not only allow for more effective allocation but could also reduce the risk of understating cases in areas with high population density.

## Discussion

The current standard of population density agnostic per capita reporting has the potential to simultaneously induce a dangerous sense of false security while accelerating infection in economic nerve centers. The contrast among the heatmaps and subsequent prospective analysis of tests and cases, unveils the scale of testing disparity which can derail the fragile path to societal normalcy and economic recovery as we *march* toward reopening the economy, after having brought it to a screeching halt in March. If we are not thoughtful in our approach to testing, this fragile bridge could collapse leading to a prolonged recession, or dare we say, a depression^9,10^.

On a positive note, fixing this is not intractable. Heatmaps of retail and payroll activity are unsurprisingly similar to population density. This is where the innate intertwining of public health and economic wellbeing around the “location, location, location” axis can be synergistic. By simply adjusting the distribution of testing capacity to also account for population density, we can improve monitoring and response to blunt the speed and spread of the virus while also safeguarding both retail activity and the economic nerve centers across the country.

## Data Availability

All study data were publicly available links cited in references.

## Author Contributions

The manuscript was written through contributions of all authors. All authors have given approval to the final version of the manuscript.

## Notes

### Competing Interest Statement

The authors have declared no competing interest.

### Funding Statement

No external funding was received.

### Author Declarations

All study data were publicly available thereby obviating institutional review board approval.

### Summary of Updates

Minor edits for ease in readability and additional references.

